# Shared genetic and neuroimmune architecture links type 1 diabetes with neurocognitive traits

**DOI:** 10.1101/2025.09.14.25335719

**Authors:** Priscilla Saarah, Zehra A. Syeda, Ziang Xu, Yikai Dong, Annie Jiang, Michelle Shanguayhia, Sourav Roy, Biqing Zhu, Le Zhang, Andrew T. Dewan, Samira Asgari, David A. Alagpulinsa

**Affiliations:** Yale Center for Molecular & Systems Metabolism, Yale University School of Medicine, New Haven, CT, USA; Department of Comparative Medicine, Yale University School of Medicine, New Haven, CT, USA; Program of Computational Biology and Bioinformatics, Yale University, New Haven, CT 06510, USA; Department of Neurology, Yale University School of Medicine, New Haven, CT, USA; Department of Neuroscience, Yale University School of Medicine, New Haven, CT, USA; Department of Chronic Disease Epidemiology, Yale School of Public Health, New Haven, CT, USA; Center for Perinatal, Pediatric and Environmental Epidemiology, Yale School of Public Health, New Haven, CT, USA; Institute for Genomic Health, Icahn School of Medicine at Mount Sinai, New York, NY, USA; Department of Genetics and Genomic Sciences, Icahn School of Medicine at Mount Sinai, New York, NY, USA

## Abstract

Type 1 diabetes (T1D), particularly with childhood onset, is associated with altered cognitive traits and neuropsychiatric risk, yet the biological bases remain unclear. Here, we integrate genome-wide association variants with single-cell epigenomic profiles and show that T1D heritability is enriched in accessible chromatin of human brain-resident cells, notably microglia, across neurodevelopment into adulthood. FDR-corrected cross-trait genetic correlation analyses revealed negative correlations of T1D with intelligence, executive function, and bipolar disorder, and a positive correlation with myasthenia gravis. Joint association analyses identified pleiotropic loci influencing both T1D and neurocognitive traits, including the neurogenomic hub 17q21.31. FDR-corrected Mendelian randomization further demonstrated protective effects of educational attainment, intelligence, Alzheimer’s disease, and bipolar disorder on T1D risk, while multiple sclerosis, myasthenia gravis, obsessive-compulsive disorder, short sleep duration, and attention-deficit/hyperactivity disorder increased T1D risk. In the reverse direction, T1D liability was associated with increased risk of myasthenia gravis and migraine, and reduced risk of schizophrenia and bipolar disorder. Genetic regulation of several brain- and immune-expressed genes, notably within 17q21.31, jointly influences T1D and neurocognitive traits, with some showing differential expression in disease-affected versus control tissue. Together, these findings highlight pleiotropic genetic and neuroimmune mechanisms that link T1D with both cognition and neuropsychiatric disease risk.

## Introduction

Type 1 diabetes (T1D) is characterized by T cell-mediated destruction of insulin-producing pancreatic beta cells, necessitating lifelong dependence on insulin therapy to control glycemia. Despite advances in management, people with T1D lose more than a decade of life expectancy [1,2] and over two decades of healthy life [3] compared with the general population.

Cognitive deficits—which are antecedents and characteristic symptoms of neuropsychiatric disorders—are consistently more prevalent in people with T1D than in the general population [4,5]. Childhood-onset T1D is particularly associated with cognitive deficits, including reduced working memory, executive function, and performance IQ [6–9]. Children with T1D often underachieve academically compared to their peers [10–12], with even poorer outcomes in those with co-occurring psychiatric disorders [10]. Neuroimaging studies support these findings, showing reduced gray and white matter volumes in children with T1D [13–15]. Adults with T1D also exhibit accelerated cognitive decline [16,17] and smaller total brain volumes [17]. Epidemiological studies further demonstrate higher rates of psychiatric disorders in people with T1D compared to the general population [12,18–20].

Because hyperglycemia is the defining pathology of T1D, research has emphasized its effects on brain development and function, identifying correlations between poor glycemic control and cognitive or psychiatric comorbidities [9,21–24]. This view has reinforced the idea that T1D-associated neurocognitive deficits arise primarily as complications of dysglycemia [9,15,25–28]. However, both T1D [29] and cognitive or neuropsychiatric disorders [30] are highly heritable—up to ∼50% and 80%, respectively. Despite this substantial heritability, the extent to which shared genetic architecture contributes to their comorbidity remains largely unexplored.

Observational studies are prone to confounding by sociodemographic factors and reverse causality—for example, impaired executive function may reduce adherence to glycemic control and exacerbate disease outcomes. Moreover, because T1D is frequently diagnosed during childhood or adolescence—a critical window for neurodevelopment [25–28,31]—clarifying whether shared genetic mechanisms jointly influence T1D and neurodevelopmental outcomes is essential.

In this study, we systematically investigate the shared genetic and cellular architecture between T1D and neurocognitive traits. By integrating genome-wide association data with single-cell epigenomic annotations, we demonstrate that T1D heritability is significantly enriched in accessible chromatin regions of brain-resident cells, particularly microglia. We further identify pleiotropic loci, genes, and genetically regulated expression patterns that converge on neuroimmune pathways, pointing to a shared genetic basis linking T1D with neurocognitive outcomes. Notably, these include the 17q21.31 locus—previously implicated in neurodevelopment and psychiatric disorders—which emerges as a key genomic hub of T1D–neurocognitive convergence.

## Results

### The heritability of T1D is enriched in brain-resident cells across development

Genetic variants associated with complex traits and diseases often localize to regulatory regions where they act in a cell type–specific manner. To test whether T1D risk variants are enriched in regulatory landscapes of brain-resident cells, we applied stratified linkage disequilibrium score regression (S-LDSC) [32] to integrate fine-mapped T1D GWAS signals (18,942 T1D cases, 520,580 individuals) [33] with single-nucleus ATAC-seq (snATAC-seq) profiles of human cortex spanning prenatal to adult stages [34]. Enrichment was defined as the proportion of T1D SNP-heritability explained by a given chromatin annotation divided by the proportion of SNPs in that annotation; values above 1 indicate more heritability than expected by chance [32,35], with significance assessed using one-sided S-LDSC P-values. This analysis revealed temporally dynamic and cell type–specific enrichment of T1D heritability, with the strongest and most consistent signal enrichment in microglia (e.g., prenatal enrichment = 5.04, z = 1.69, one-sided P = 0.045; adult enrichment = 4.36, z = 1.74, P = 0.041) (**Fig. 1a**). Astrocytes also showed nominal enrichment in the prenatal stage, whereas other glial and neuronal lineages showed little signal.

**Figure 1.**
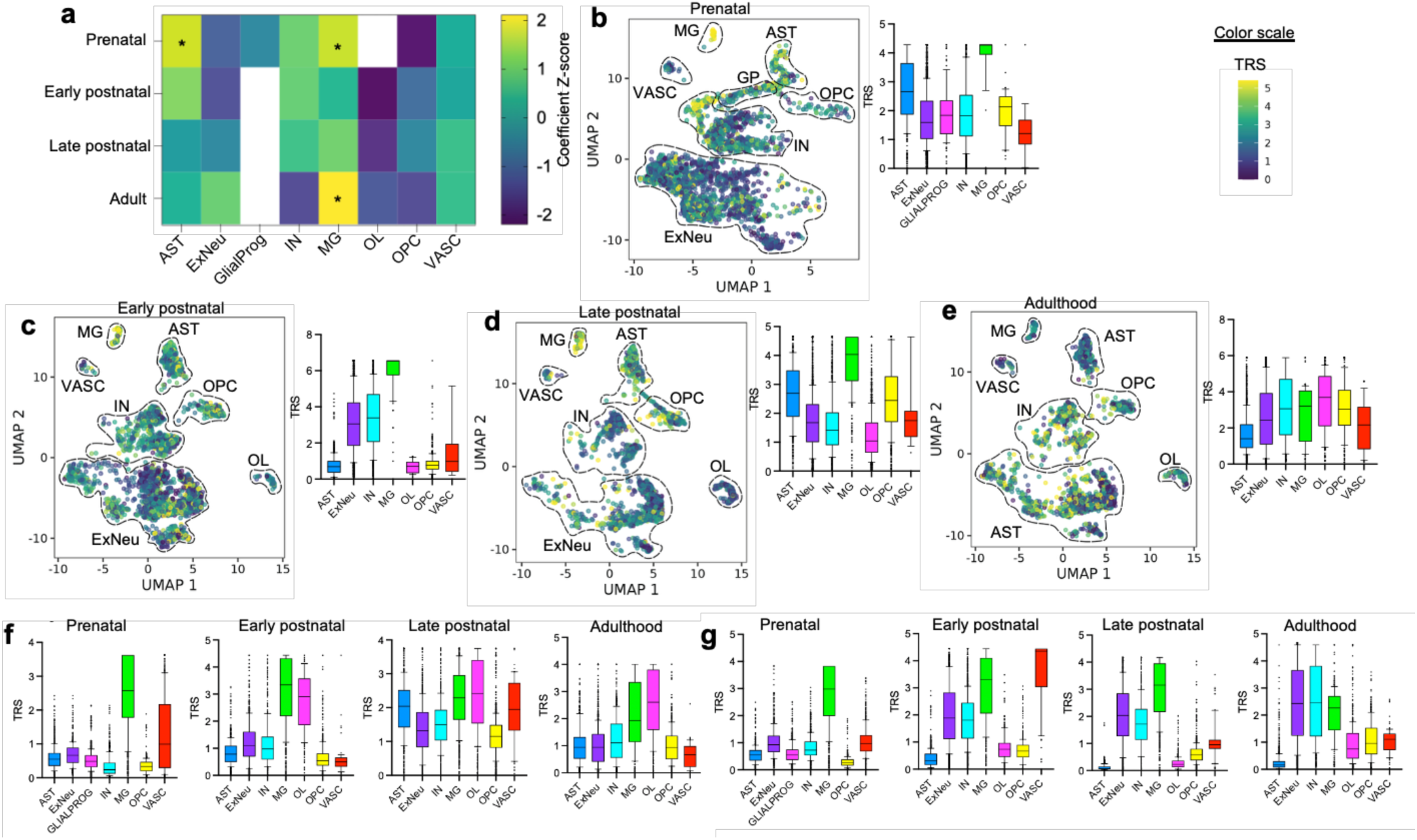
Enrichment of T1D heritability in brain-resident cell types across neurodevelopment. (**a**) Stratified LD score regression (S-LDSC) of fine-mapped T1D GWAS variants in accessible chromatin of brain cell types—astrocytes (AST), excitatory neurons (ExNeu), glial progenitors (GLIALPROG), inhibitory neurons (IN), microglia (MG), oligodendrocyte progenitor cells (OPC), and vascular cells (VASC)—across four neurodevelopmental stages (prenatal, early postnatal, late postnatal, and adult), profiled by single-nucleus ATAC-seq (snATAC-seq). The heatmap displays enrichment scores (observed h² proportion relative to SNP proportion), with asterisks indicating nominal significance (one-sided P < 0.05). (**b–e**) SCAVENGE trait-relevance scores (TRS) for T1D fine-mapped variants in snATAC-seq data across prenatal (**b)**, early postnatal (**c**), late postnatal (**d**), and adult (**e**) stages. (**f**) SCAVENGE TRS based on Alzheimer’s disease GWAS-significant variants. (**g**) SCAVENGE TRS based on bipolar disorder GWAS-significant variants. Box plots show the interquartile range (IQR; box), median (center line), and 1.5× IQR (whiskers).

Because S-LDSC aggregates across nuclei and may under-detect transient or sparse signals in heterogeneous single-cell data, we next applied SCAVENGE (Single Cell Analysis of Variant Enrichment through Network propagation of Genomic annotations), a network-propagation framework that assigns trait-relevance scores (TRS) at single-cell resolution [36], SCAVENGE confirmed persistent enrichment in microglia across all stages (median TRS >3.0–6.5) and further highlighted context-specific signals in excitatory and inhibitory neurons (early postnatal) and oligodendrocyte lineage cells (adulthood) (Fig. 1b–e). To contextualize these results, we compared enrichment patterns for Alzheimer’s disease (AD) and bipolar disorder (BPD), two traits with known brain-cell architectures [37]. As expected, AD signals localized predominantly to microglia, whereas BPD showed enrichment in neurons during adulthood and in microglia at earlier stages (**Fig. 1f, g**).

Together, these complementary analyses converge on microglia as the primary mediators of T1D genetic liability in the brain, with additional stage-specific contributions from other neural and glial populations.

### The genetic architecture of T1D overlaps with neurocognitive traits

Given the enrichment of T1D heritability in brain-resident cells, we next tested for genome-wide genetic overlap with neurocognitive traits using cross-trait linkage disequilibrium score regression (LDSC) across 21 cognitive, psychiatric, neurological, and sleep-related traits (**Supplementary Table 1**). After FDR correction, T1D showed significant negative genetic correlations with intelligence (*r*_g_ = –0.09, SE = 0.02, p = 7.8×10⁻⁵, q = 5.4×10⁻⁴), bipolar disorder (*r*_g_ = –0.11, SE = 0.03, p = 6.4×10⁻⁵, q = 5.4×10⁻⁴), and executive function (*r*_g_ = –0.08, SE = 0.03, p = 0.0013, q = 0.0068), and positive correlations with myasthenia gravis (*r*_g_ = 0.37, SE = 0.08, p = 3×10⁻⁶, q = 7.0×10⁻⁵) and migraine (*r*_g_ = 0.10, SE = 0.04, p = 0.0112, q = 0.0470). Nominal associations (p < 0.05 but q > 0.05) included negative correlations with educational attainment (*r*_g_ = –0.06, SE = 0.03, p = 0.0200, q = 0.0700) and autism spectrum disorder (*r*_g_ = –0.11, SE = 0.05, p = 0.0271, q = 0.0813), and positive correlations with insomnia (*r*_g_ = 0.07, SE = 0.03, p = 0.0345, q = 0.0906) and multiple sclerosis (*r*_g_ = 0.20, SE = 0.10, p = 0.0397, q = 0.0926). Other traits, including Alzheimer’s disease, schizophrenia, OCD, ischemic stroke, and neuroticism, showed no evidence of genetic correlation with T1D (**Fig. 2a; Supplementary Table 2**).

**Figure 2.**
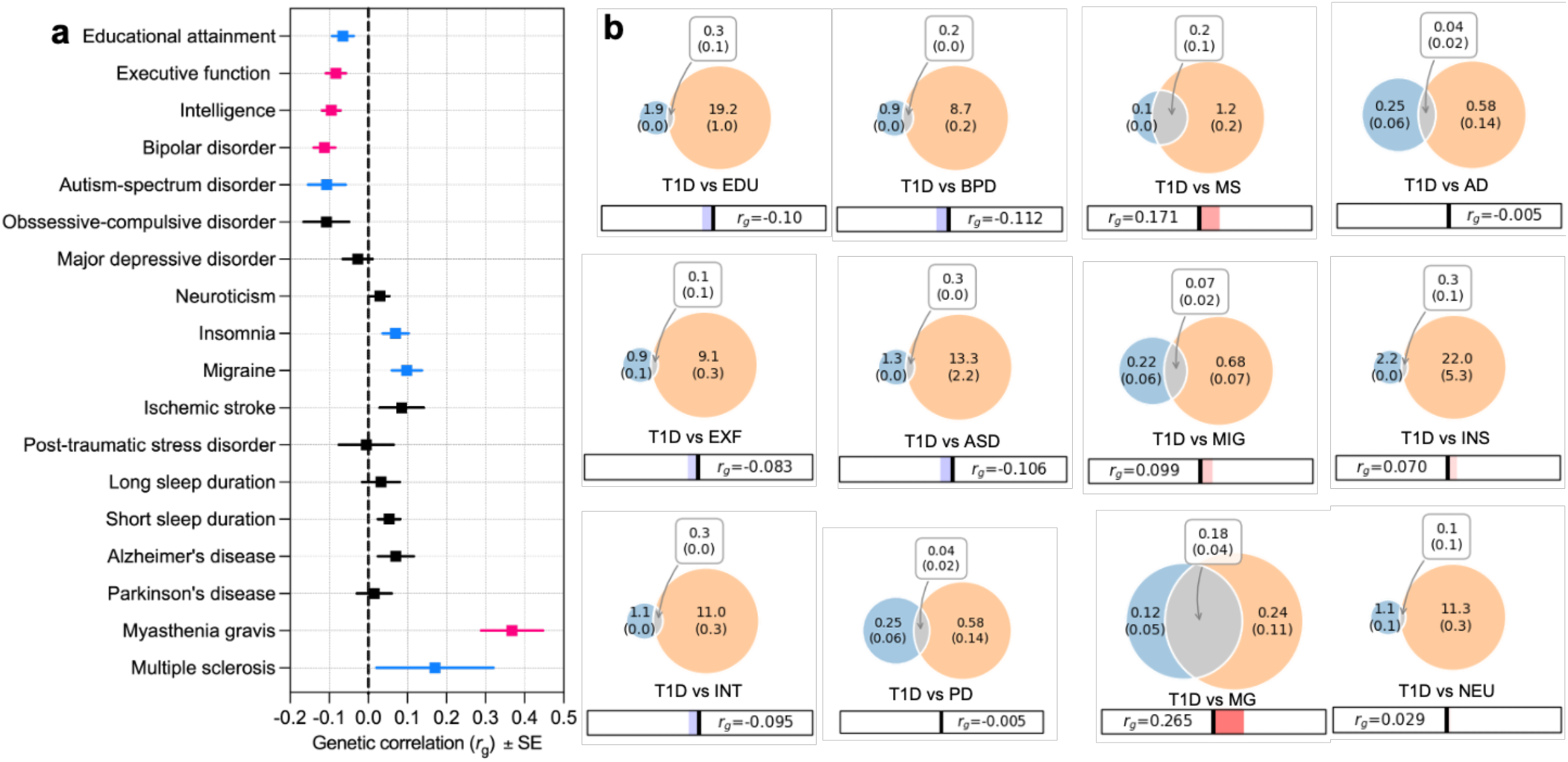
Genetic correlations and polygenic overlap between T1D and neurocognitive traits. (**a**) Cross-trait linkage disequilibrium score regression (LDSC) estimates of genome-wide genetic correlations (*r*_g_) between T1D and cognitive, psychiatric, and neurological traits. Squares indicate point estimates, with horizontal bars showing standard errors. Red/pink markers denote correlations significant after false discovery rate (FDR) correction (q < 0.05), while blue markers denote nominal associations (p < 0.05, q ≥ 0.05). (**b**) MiXeR bivariate causal mixture models estimating the number of trait-influencing variants for each phenotype (orange) and for T1D (blue), with the overlapping area representing shared variants. Numbers inside circles indicate estimated polygenicity (s.e.), and numbers in the overlap indicate the number of shared causal variants. *r*_g_ values from LDSC are shown below each panel.

To further dissect the genetic overlap between T1D and neurocognitive traits, we applied MiXeR, which estimates polygenicity and quantifies the number of causal variants shared between traits. Unlike LDSC, which summarizes genome-wide correlation as an average effect direction, MiXeR captures pleiotropy irrespective of direction. MiXeR revealed extensive variant sharing between T1D and neurocognitive traits (**Fig. 2b**). MiXeR showed that T1D is moderately polygenic (∼22,000 variants) and shares hundreds of causal variants with nearly all examined neurocognitive traits. The strongest overlap was with myasthenia gravis, consistent with their shared autoimmune basis, but notable overlap was also observed with intelligence and educational attainment. Importantly, these overlaps persisted despite negative genetic correlations, indicating that many of the same variants influence both T1D and cognition but with opposite effect directions.

Overall, the LDSC and MiXeR analyses provide complementary perspectives: LDSC highlights the direction of genome-wide associations, while MiXeR underscores the pervasiveness of shared causal variants irrespective of direction, supporting a model in which common brain–immune pathways exert pleiotropic influences across autoimmune, cognitive, and psychiatric outcomes.

### Conjunctional false discovery analysis identifies loci jointly influencing T1D and neurocognitive traits

To resolve locus-level overlap, we applied conjunctional false discovery rate (conjFDR) analysis, which leverages cross-trait enrichment to identify pleiotropic loci beyond conventional genome-wide significance thresholds [38,39]. Across trait pairs, the number of loci jointly associated with T1D and neurocognitive traits ranged from fewer than 10 to more than 100, with particularly extensive overlap for neuroautoimmune disorders such as multiple sclerosis (109 loci) and myasthenia gravis (69 loci) (**Fig. 3a-h; Supplementary Table 3**). Substantial sharing was also evident for cognitive traits, including intelligence (63 loci) and educational attainment (45 loci), and for psychiatric and neurodegenerative conditions such as Alzheimer’s disease (29 loci), bipolar disorder (13 loci), and Parkinson’s disease (9 loci).

**Figure 3.**
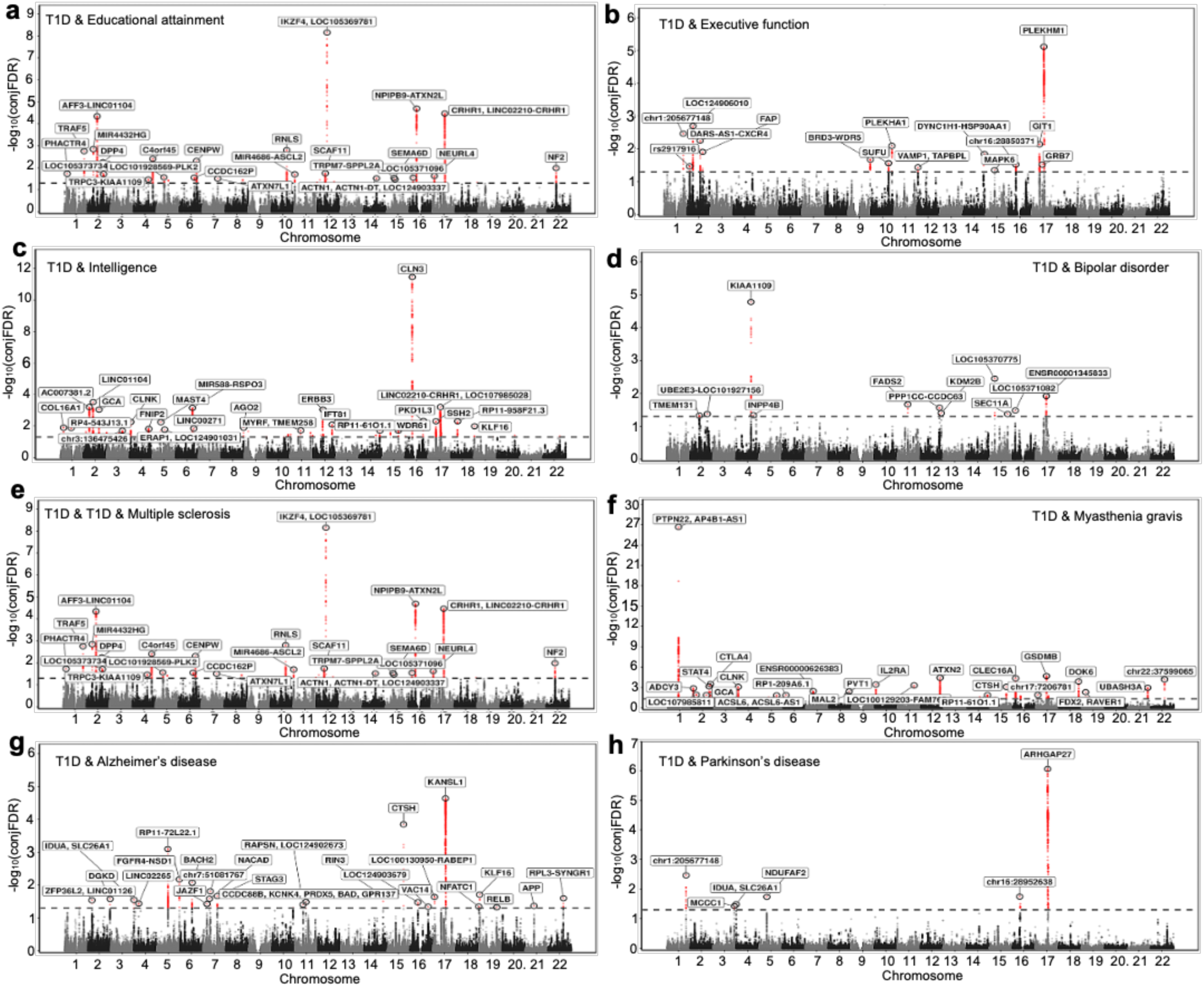
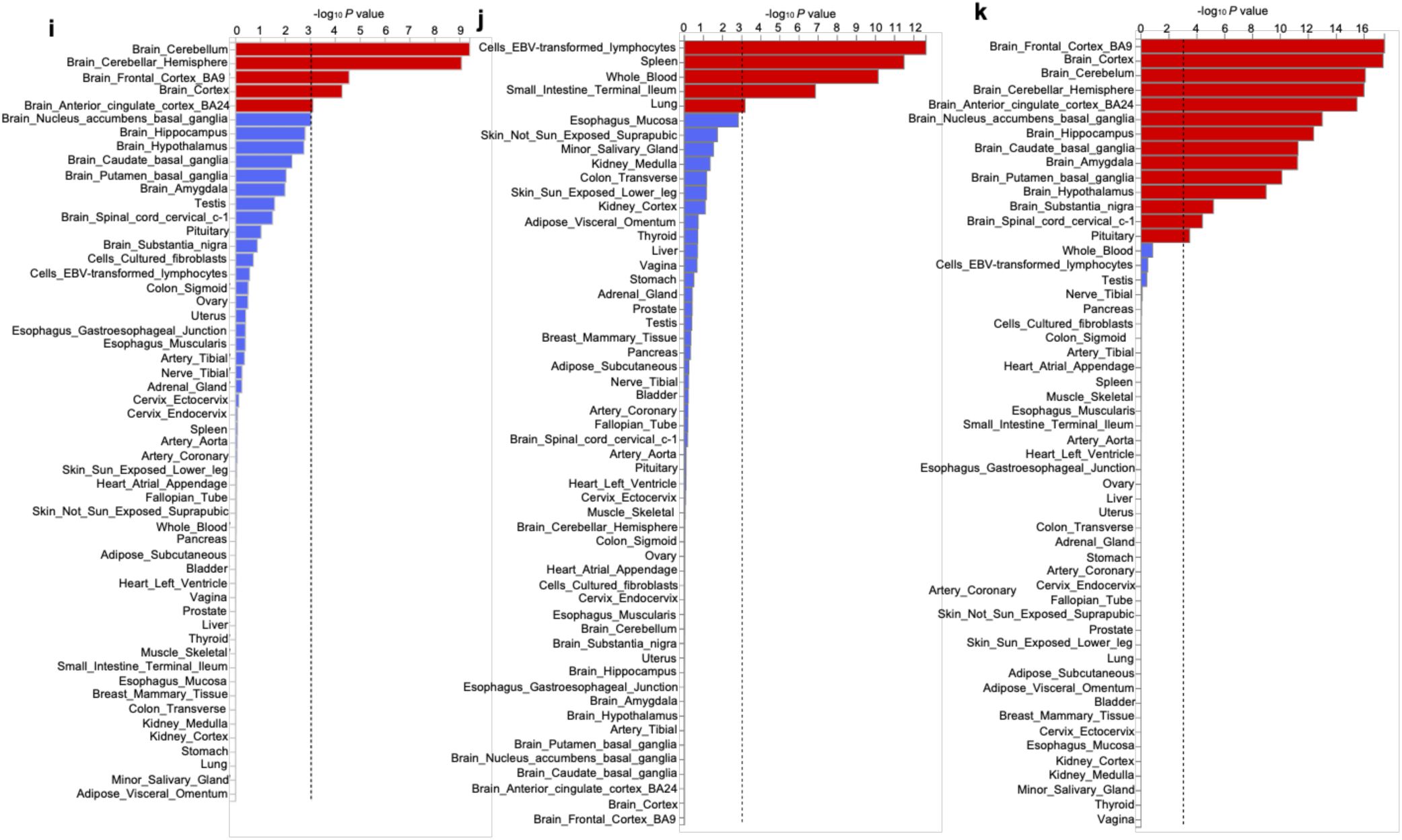
Conjunctional false discovery rate (conjFDR) and tissue enrichment analyses of loci jointly associated with T1D and neurocognitive traits. (**a–h**) ConjFDR Manhattan plots showing loci jointly associated with T1D and educational attainment (**a**), executive function (**b)**, intelligence (**c**), bipolar disorder (**d**), multiple sclerosis (**e**), myasthenia gravis (**f**), Alzheimer’s disease (**g**), and Parkinson’s disease (**h**). Red points denote loci surpassing the conjFDR significance threshold (conjFDR < 0.05), annotated with nearest genes. (**i–k**) Tissue enrichment of pleiotropic loci jointly associated with T1D and educational attainment (**i**), multiple sclerosis (**j**), and bipolar disorder (**k**). Bars represent – log10(P) values for enrichment across GTEx tissues, with the dashed line indicating nominal significance.

Among the shared loci, the **17q21.31** region emerged as a pleiotropic hotspot, jointly associated with T1D and multiple neurocognitive traits. This structurally polymorphic region contains *MAPT*, *KANSL1*, *CRHR1*, and *LRRC37A*—genes implicated in neurodevelopment, neurodegeneration, and immune regulation—highlighting 17q21.31 as a candidate neuroimmune hub. Notably, this locus has been associated previously with cognitive phenotypes and various neuropsychiatric or neurodevelopmental disorders [40–43].

To assess biological context, we examined tissue-specific enrichment of pleiotropic loci. We focused on three representative pairs spanning cognitive, autoimmune, and psychiatric axes: educational attainment, multiple sclerosis, and bipolar disorder (**Fig. 3i-k**). Shared loci between T1D and educational attainment (45 loci) were enriched in brain regions including the cerebellum, frontal cortex, and anterior cingulate cortex. Shared loci between T1D and multiple sclerosis (109 loci) were enriched in immune-related tissues such as EBV-transformed lymphocytes, spleen, whole blood, and small intestine. Shared loci between T1D and bipolar disorder (13 loci) were enriched in cortical and subcortical brain regions including frontal cortex, hippocampus, putamen, and amygdala.

These results show that pleiotropic loci linking T1D with neurocognitive traits are distributed in biologically coherent patterns: enriched in brain tissues for cognitive and psychiatric traits, and in immune tissues for neuroautoimmune traits, underscoring the dual brain–immune architecture of T1D pleiotropy.

### Bidirectional Mendelian randomization identifies directional and reciprocal associations between T1D and neurocognitive traits

To investigate potential causal relationships, we conducted bidirectional two-sample Mendelian randomization (MR) using inverse variance weighted (IVW) as the primary estimator, complemented by weighted median and MR-Egger models to assess robustness and pleiotropy. False discovery rate (FDR) correction was applied across all 42 IVW tests.

When modeling genetic liability to T1D as the exposure (**Fig. 4a; Supplementary Table 4**), we observed FDR-significant associations with increased risk of myasthenia gravis (OR = 1.20, 95% CI: 1.12–1.28, P = 7.0×10⁻⁸, q = 3×10⁻⁶) and migraine (OR = 1.02, 95% CI: 1.01–1.03, P = 1.8×10⁻³, q = 0.010). Conversely, T1D liability was associated with reduced risk of schizophrenia (OR = 0.98, 95% CI: 0.96–0.99, P = 7.6×10⁻³, q = 0.030) and bipolar disorder (OR = 0.98, 95% CI: 0.96–0.99, P = 8.9×10⁻³, q = 0.032). This inverse association with schizophrenia is consistent with a recent analysis of individual-level data, which reported reduced schizophrenia risk among people with T1D in both observational and MR frameworks, although the MR associations weakened after multiple-testing correction [44]. Several additional associations reached nominal significance but did not survive FDR correction, including ischemic stroke (OR = 1.01, 95% CI: 1.00–1.02, P = 0.029, q = 0.24), Parkinson’s disease (OR = 0.97, 95% CI: 0.95–1.00, P = 0.048, q = 0.08), and amyotrophic lateral sclerosis (ALS; OR = 0.98, 95% CI: 0.96–1.00, P = 0.020, q = 0.07). Our nominal finding for Parkinson’s disease liability aligns with prior MR evidence suggesting a bidirectional relationship with T1D [45,46].

**Figure 4.**
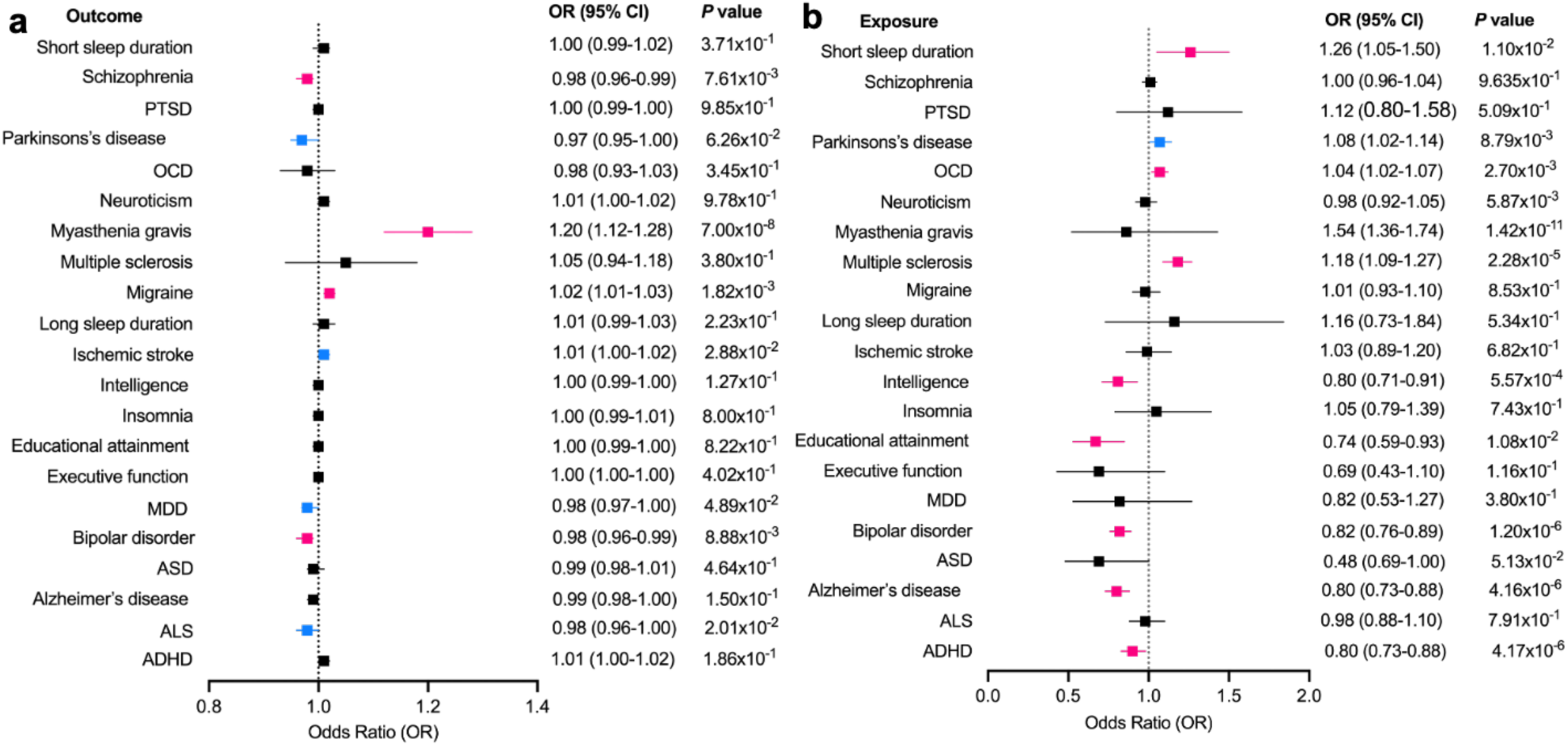
Bidirectional Mendelian randomization (MR) analysis of T1D and neurocognitive traits. (a) Effects of genetic liability to T1D on risk of neurocognitive, psychiatric, and neurological traits. (**b**) Effects of genetic liability to neurocognitive, psychiatric, and neurological traits on risk of T1D. Odds ratios (squares) with 95% confidence intervals (horizontal lines) are shown from inverse variance weighted (IVW) models. Red/pink markers denote associations significant after false discovery rate (FDR) correction (q < 0.05), while blue markers denote nominal associations (p < 0.05, q ≥ 0.05). Results from weighted median and MR-Egger models are provided in Supplementary Table 4.

When modeling neurocognitive traits as exposures (**Fig. 4b; Supplementary Table 4**), higher liability to multiple sclerosis (OR = 1.18, 95% CI: 1.09–1.27, P = 2.3×10⁻⁵, q = 2×10⁻⁴), myasthenia gravis (OR = 1.22, 95% CI: 1.12–1.33, P = 5.6×10⁻⁶, q = 4×10⁻⁵), obsessive–compulsive disorder (OR = 1.07, 95% CI: 1.02–1.12, P = 3.7×10⁻³, q = 0.016), short sleep duration (OR = 1.26, 95% CI: 1.05–1.50, P = 0.011, q = 0.036), and ADHD (OR = 0.90, 95% CI: 0.83–0.98, P = 0.016, q = 0.048) was associated with increased T1D risk. In contrast, liability to educational attainment (OR = 0.67, 95% CI: 0.53–0.85, P = 8.2×10⁻⁴, q = 0.006), intelligence (OR = 0.81, 95% CI: 0.71–0.93, P = 2.0×10⁻³, q = 0.010), Alzheimer’s disease (OR = 0.80, 95% CI: 0.73–0.88, P = 4.2×10⁻⁶, q = 5×10⁻⁵), and bipolar disorder (OR = 0.82, 95% CI: 0.76–0.89, P = 1.2×10⁻⁶, q = 2×10⁻⁵) was associated with reduced risk of T1D. Parkinson’s disease also showed a nominal association with increased T1D risk (OR = 1.07, 95% CI: 1.01–1.14, P = 0.019, q = 0.08), but this did not pass FDR correction.

Collectively, these analyses reveal both directional and reciprocal associations between T1D and neurocognitive traits.

### Shared gene regulation in brain and immune cells links T1D and neurocognitive traits

To test whether shared genetic risk between T1D and neurocognitive traits is mediated through gene expression, we performed summary-based Mendelian randomization with HEIDI filtering (SMR/HEIDI) across brain tissues, single-nucleus brain cell types, and immune populations. These analyses revealed that shared regulatory signals were concentrated at 17q21.31 and extended across additional loci (**Fig. 5; Supplementary Table 5**).

**Figure 5.**
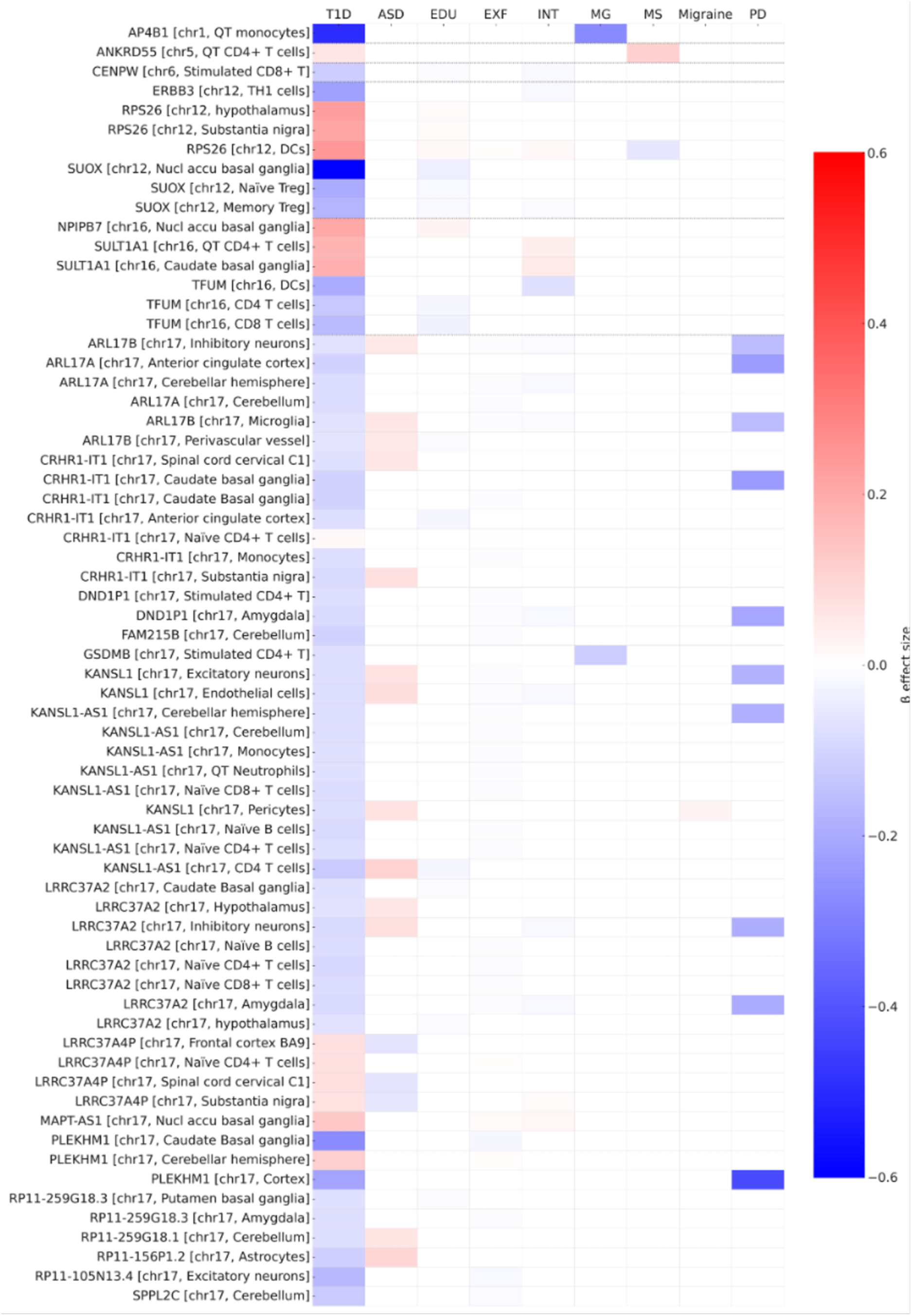
Shared gene regulation in brain and immune cells links T1D with neurocognitive traits. Heatmap of genes identified by summary-based Mendelian randomization with HEIDI filtering (SMR/HEIDI), showing eQTLs with pleiotropic associations between T1D and at least one neurocognitive trait. Genes are ordered by chromosome, with the tissue or cell type in which the regulatory effect was detected shown in brackets. Columns represent traits: autism spectrum disorder (ASD), educational attainment (EDU), executive function (EXF), intelligence (INT), myasthenia gravis (MG), multiple sclerosis (MS), migraine, and Parkinson’s disease (PD). Color scale represents the direction and magnitude of β effect sizes, with red indicating higher gene expression associated with increased disease risk (or trait liability) and blue indicating higher gene expression associated with reduced risk. Vertical dotted lines denote chromosome boundaries across genes. Only genes passing FDR-corrected significance thresholds for SMR and with HEIDI P > 0.05 are shown.

At 17q21.31, multiple transcripts demonstrated pleiotropic associations across brain and immune contexts. Increased expression of CRHR1-IT1 and LRRC37A2 in substantia nigra, hypothalamus, amygdala, inhibitory neurons, and T-cell subsets was associated with reduced T1D risk but higher ASD risk, while overlapping with EDU, EXF, and PD. In contrast, LRRC37A4P and MAPT-AS1 expression correlated with elevated T1D risk and higher INT/EXF, with LRRC37A4P also linked to ASD, EDU, MG, and PD. Other transcripts—including KANSL1-AS1, ARL17A/B, RP11-259G18 isoforms, PLEKHM1, SPPL2C, FAM215B, DND1P1, and KANSL1—generally showed reduced T1D risk accompanied by lower INT/EXF/EDU, with additional trait-specific associations (e.g., KANSL1-AS1 with ASD, EDU, EXF, INT, MG, MS, PD; ARL17B with ASD, EDU, EXF, INT, PD, migraine). These effects were detected across diverse brain-resident cells (microglia, neurons, astrocytes, pericytes, endothelial cells) and immune states (T- and B-cell subsets), underscoring 17q21.31 as a central neuroimmune regulatory hub.

Beyond 17q21.31, additional loci reinforced this brain–immune convergence. On chromosome 12, SUOX expression in nucleus accumbens and regulatory T cells was linked to reduced T1D and lower EDU/INT/EXF, whereas RPS26 expression in substantia nigra, hypothalamus, and dendritic cells was associated with elevated T1D, higher EDU/INT/EXF, and lower MS. On chromosome 16, NPIPB7 in nucleus accumbens linked higher T1D with higher EDU, while SULT1A1 in caudate and T cells linked higher T1D with higher INT/EDU. TFUM in CD4+, CD8+, and dendritic cells associated with lower T1D and lower EDU/INT/EXF, overlapping with MG, MS, and PD. Other notable associations included CENPW (chr6; stimulated CD8+ T cells) linking reduced T1D with lower EDU/INT/EXF; ANKRD55 (chr5; CD4+ T cells) linking elevated T1D with MS and EDU/INT; ERBB3 (chr12; TH1 cells) linking lower T1D with lower INT; AP4B1 (chr1; monocytes) linking lower T1D with lower MG/INT/EXF/MS; and GSDMB (chr17; CD4+ T cells) linking lower T1D with lower MG/EDU/EXF.

Overall, these results indicate that shared liability between T1D and neurocognitive traits is mediated by pleiotropic gene regulation across brain and immune cells. While LDSC highlighted negative genome-wide correlations between T1D and cognition alongside paradoxical positive correlation between ASD and cognition, eQTL integration clarified this architecture: T1D and cognition typically move in the same direction, whereas ASD diverges. Gene regulation—anchored at 17q21.31 and extending to loci on chromosomes 1, 5, 6, 12, and 16—thus emerges as a key mechanistic axis linking autoimmunity, neurodevelopment, and cognition.

### Genes regulated by eQTLs that jointly influence T1D and neurocognitive traits exhibit differential expression between disease-affected and control brain samples

Our Mendelian randomization (MR) analyses provided **nominal evidence** of a bidirectional association between T1D and Parkinson’s disease (PD), with the strongest effect consistent with PD liability increasing T1D risk. These associations did not withstand FDR correction, but convergent signals from conjunctional FDR and SMR/HEIDI analyses— particularly within the 17q21.31 locus—highlighted shared eQTLs influencing both T1D and neurocognitive traits. Moreover, prior independent studies have reported similar causal relationships between T1D and PD [45,46], lending external support to this observed connection. To evaluate whether these genetically predicted regulatory effects manifest in human disease, we interrogated our previously published single-nucleus RNA-sequencing (snRNA-seq) and proteomic datasets from PD and control prefrontal cortex [47]. Specifically, we examined genes whose genetically predicted expression in bulk brain regions and single-cell populations was associated with T1D risk.

Analysis of genes whose genetically predicted expression in cortex is associated with T1D risk revealed that several 17q21.31 transcripts protective for T1D (e.g., *LRRC37A2, KANSL1-AS1, CRHR1, ARL17A,* and *PLEKHM1*) were significantly downregulated in PD neurons (**Fig. 6a**). This expression pattern is concordant both with their protective SMR effect on T1D and with the MR inference that PD liability increases T1D risk through suppression of protective genes. In contrast, *CCDC88B*, whose increased expression is predicted to elevate T1D risk, was upregulated in PD glia, again consistent with its genetically inferred effect direction.

**Figure 6.**
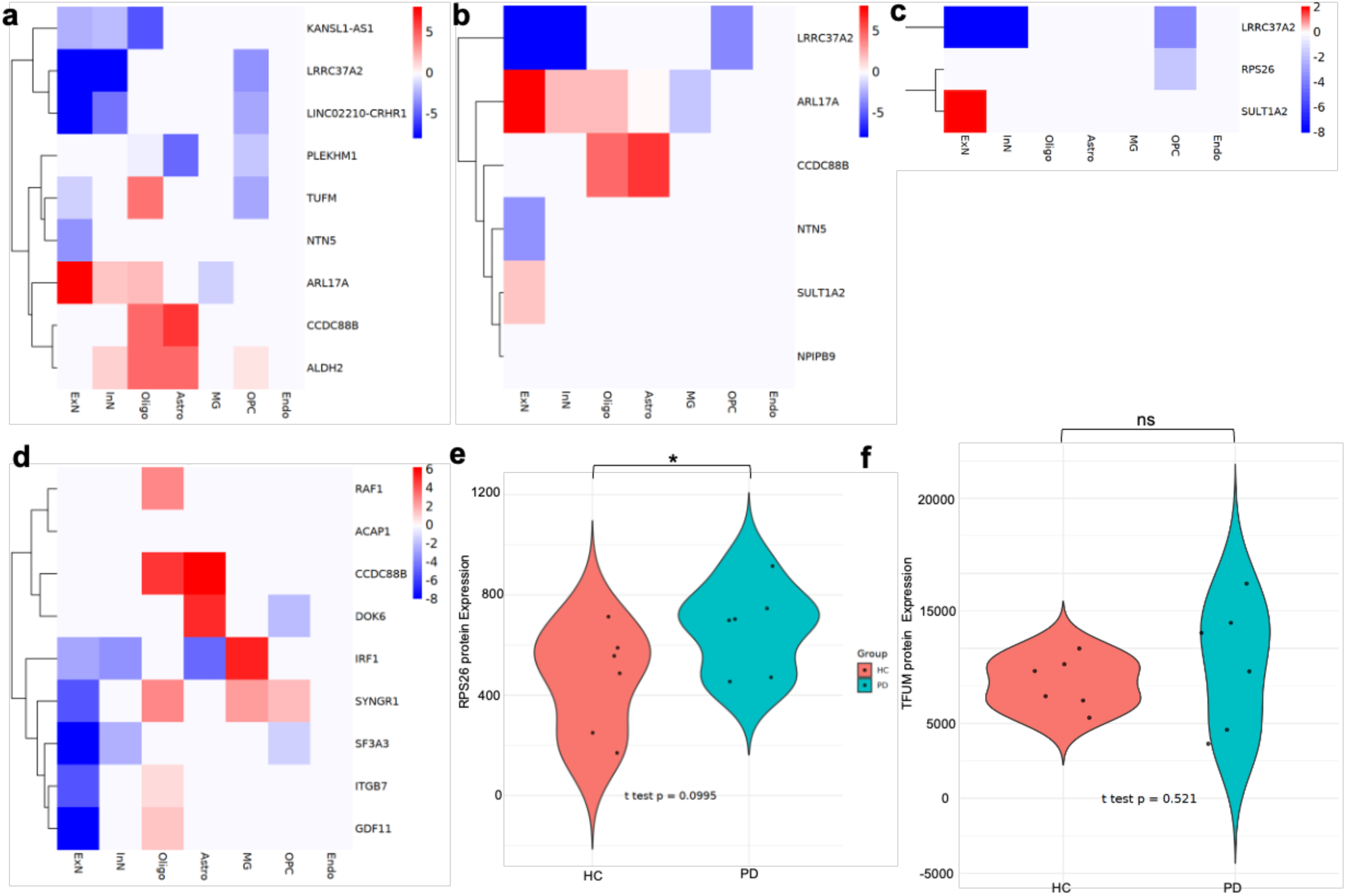
Differential transcriptomic and proteomic expression of SMR/HEIDI-identified genes in Parkinson’s disease brain tissue. Genes whose genetically predicted expression in cortex (**a**), frontal cortex BA9 (**b**), substantia nigra (**c**), or microglia (**d**) were associated with T1D risk in SMR/HEIDI analyses were assessed for differential expression in single-nucleus RNA-seq datasets from Parkinson’s disease (PD) and control prefrontal cortex. Each heatmap depicts normalized transcript expression (z-score) for PD versus control samples, with rows corresponding to genes and columns to individual samples. Statistical significance was determined using DESeq2 with covariate adjustment for age, sex, and postmortem interval; only genes with adjusted *P* < 0.05 are shown. (**e–f**) Differential protein abundance of SMR-identified genes with brain-specific eQTL associations with T1D, quantified using label-free mass spectrometry in the same PD and control cohort. Bars represent mean normalized protein abundance ± standard error. Significance was determined using moderated *t*-tests with Benjamini–Hochberg correction. A relaxed significance threshold of *P* < 0.1 was used due to small sample size; proteins marked with an asterisk (*) reached *P* < 1.

Genes whose genetically predicted expression in BA9 is associated with T1D risk displayed similar relationships (**Fig. 6b**). Protective transcripts such as *LRRC37A2* and *ARL17A* were reduced in PD neurons, while risk-promoting genes including *CCDC88B* and *SULT1A2* were upregulated in glial populations. These findings reinforce that PD liability enhances T1D risk via downregulation of protective and upregulation of risk genes.

In substantia nigra, *LRRC37A2* and *RPS26* transcripts were reduced in neurons, while *SULT1A2* expression was increased in non-neuronal populations (**Fig. 6c**). Notably, although *RPS26* showed transcript downregulation, our proteomics analysis demonstrated significant upregulation of RPS26 protein in PD cortex (**Fig. 6e–f**). This result is fully consistent with the SMR and MR predictions that higher *RPS26* expression increases T1D risk and highlights the importance of post-transcriptional regulation.

Interrogation of microglial genes whose genetically predicted expression is associated with T1D revealed a coordinated activation pattern in PD (**Fig. 6d**). Risk-promoting genes such as *RAF1, ACAP1,* and *DOK6* were upregulated, while protective genes including *IRF1, SYNGR1, SFTA3, ITGB7,* and *GDF11* were downregulated. This directionality is fully consistent with the MR inference that PD risk exacerbates T1D risk through immune-mediated dysregulation.

Proteomic profiling confirmed significant upregulation of RPS26 in PD cortex (**Fig. 6e–f**), directly validating SMR and MR predictions that increased RPS26 expression promotes T1D risk. Other SMR-prioritized genes such as *TUFM* did not show significant protein changes, highlighting gene-specific differences in post-transcriptional regulation.

Together, these analyses provide direct empirical support that PD liability increases T1D risk via suppression of protective and induction of risk-promoting genes in brain regions and immune cell types. Our integration of SMR/HEIDI inference with snRNA-seq and proteomics thus reveals convergent neuroimmune pathways—particularly centered on 17q21.31 and *RPS26*—that mechanistically link PD and T1D.

## Discussion

T1D, while classically viewed as a peripheral autoimmune disease, has long been associated with cognitive dysfunction and psychiatric disease risk, particularly in those with childhood onset [12,18–20]. However, the biological basis of this comorbidity has been poorly understood. Here, through an integrative genomic approach, we uncover convergent genetic, regulatory, and cellular mechanisms linking T1D with a spectrum of neurocognitive traits. These findings support a neuroimmunogenetic model of T1D, in which liability to autoimmunity and altered neurodevelopment reflects shared molecular architecture.

Our study reveals that T1D heritability is enriched in accessible chromatin regions of brain-resident cells, particularly microglia, across prenatal and postnatal development. Microglia are central players in immune surveillance and synaptic refinement, positioning them at the nexus of neural–immune crosstalk. The enrichment of T1D genetic risk in microglia during early development suggests that genetic alterations of neuroimmune programs may influence both autoimmune susceptibility and neurodevelopmental processes before T1D onset, potentially contributing to the cognitive alterations frequently observed in patients with early-onset disease.

The genome-wide genetic correlation and polygenic overlap analyses further demonstrated that T1D shares substantial, and often inverse, genetic architecture with cognitive and psychiatric traits. Notably, we observed negative genetic correlations between T1D and educational attainment (EDU), executive function (EXF), and intelligence (INT)— traits that also exhibited causal associations with T1D in Mendelian randomization analyses. Genetic liability to higher EDU and INT reduced T1D risk, suggesting a potential neurodevelopmental resilience against autoimmunity. Conversely, T1D liability conferred increased risk for neuroimmune disorders such as myasthenia gravis and migraine, while reducing risk for schizophrenia and bipolar disorder, and showing nominal evidence for reduced risk of Parkinson’s disease. The inverse association with schizophrenia is consistent with prior MR analyses, where associations weakened after multiple-testing correction, and with individual-level data showing reduced schizophrenia risk among individuals with T1D [44]. Although our MR analyses only provided nominal evidence for associations between T1D and Parkinson’s disease, the observation that genetic predisposition to T1D trended toward reduced PD risk, while PD liability showed a nominal association with increased T1D risk is intriguing [45,46]. This asymmetry, also reported in prior studies [45,46], may reflect complex pleiotropic mechanisms where dopaminergic neurodegeneration in PD promotes systemic immune dysregulation [48], while T1D-related immune mechanisms could potentially confer resilience against neurodegeneration. In contrast, T1D and bipolar disorder demonstrate inverse causal effects in both directions— where liability to either trait reduces the risk of the other—consistent with their negative genetic correlation and polygenic overlap. Together, these bidirectional relationships exemplify how reciprocal genetic influences can shape comorbidity, risk antagonism, or protective divergence between disorders, pointing to distinct etiological axes of brain– immune interaction. The causal associations of T1D with migraine and myasthenia gravis further underscore the role of neuroimmune crosstalk, as both conditions are increasingly recognized to involve overlapping immune and neuronal regulatory pathways [49,50]. Similar nominal associations were also observed for amyotrophic lateral sclerosis (ALS), consistent with evidence that glial and immune dysfunction contribute to ALS pathogenesis [51], suggesting possible shared mechanisms with neurodegeneration that warrant further investigation.

To probe potential mechanisms, we identified numerous pleiotropic variants using conjunctional FDR and SMR/HEIDI analyses. The 17q21.31 locus emerged as a key neuroimmune hub, harboring eQTLs that regulate gene expression in both brain and immune cells that influence traits spanning T1D, cognition, and neuropsychiatric phenotypes. This region, long implicated in schizophrenia, bipolar disorder, autism, and depression [40–43], contains genes such as CRHR1, KANSL1, and MAPT with broad roles in synaptic plasticity, stress response, and neurodevelopment [42,43]. Our analyses extend this spectrum by showing that genetically predicted expression of several genes in this locus (e.g., CRHR1, KANSL1, PLEKKHM1, and LRRC37A2), as well as RPS26 on chromosome 12, is associated with risk of T1D and neurocognitive traits. Moreover, these genes exhibit disease-associated expression changes in single-nucleus RNA-seq and proteomic data from Parkinson’s disease brains, often in directions concordant with genetically predicted T1D risk. These molecular signatures support shared etiological pathways mediated through transcriptional regulation in brain-resident cells and suggest that the 17q21.31 locus may act as a convergence point for immune and cognitive processes relevant to both autoimmunity and neuropsychiatric disease.

Although LDSC analyses revealed significant negative genetic correlations between T1D and cognitive traits (EDU, INT, EXF), as well as ASD, our expression-based colocalization findings added a more nuanced layer. Specifically, most eQTLs with pleiotropic effects on T1D and EDU/INT/EXF showed expression-mediated effects in the same direction—i.e., increased gene expression was associated with both increased cognitive performance and elevated T1D risk. This appears discordant with the negative genome-wide genetic correlation and the protective MR effects of EDU and INT on T1D. However, this discrepancy reflects fundamental differences in resolution: LDSC captures the average direction of genome-wide polygenic overlap [52], whereas SMR highlights specific loci with expression-mediated pleiotropy [53]. The MR results support a causal effect of cognitive traits on reduced T1D risk, but this causal pathway likely includes mechanisms not captured by eQTL-mediated expression alone. Although ASD did not show FDR-significant causal effects, its divergent SMR patterns highlight mechanistic differences compared to other cognitive traits. These patterns underscore that genome-wide polygenic relationships, causal trait influences, and regulatory mechanisms can diverge in direction and tissue-specificity.

Collectively, the results of this study redefine our understanding of T1D by situating it within a broader neurodevelopmental and neuroimmune context. Rather than an isolated autoimmune pathology, T1D emerges as a pleiotropic trait whose genetic architecture converges with that of cognition, psychiatric traits, and neuroinflammation. This convergence is rooted in shared regulatory variation, developmentally dynamic brain–immune interactions, and pleiotropic gene expression programs spanning both central and peripheral systems.

Our study has important implications. First, it motivates deeper mechanistic investigation of microglial and glial contributions to T1D pathogenesis—particularly during early neurodevelopment. Second, it highlights potential biomarkers and therapeutic targets—such as CRHR1, LRRC37A2 and KANSL1—that may modulate both immune and neurocognitive outcomes. Third, it underscores the utility of integrative genomics in uncovering complex pleiotropy, even in traits traditionally viewed as organ-specific. Finally, it suggests that cognitive and psychiatric features in T1D may not be mere complications of dysglycemia but instead reflect a shared genetic liability that predates clinical onset.

The findings of this study are not without limitations. Although Mendelian randomization supports directional inference, sample overlap and residual pleiotropy cannot be fully excluded. Our use of UK Biobank data may introduce shared participants across some GWAS datasets, potentially biasing causal estimates. Furthermore, while we leveraged diverse eQTL resources, tissue- and context-specific regulatory dynamics remain incompletely captured, particularly in disease-relevant states. Future work integrating perturbation-based single-cell models and longitudinal cohorts will be critical to functionally validate these findings. Finally, while our analyses focused on individuals of predominantly European ancestry, the extent to which these neuroimmune mechanisms generalize across global populations remains to be determined.

In summary, this study provides genomic evidence that T1D is embedded within a broader neurocognitive and neuroimmune architecture. The convergence of genetic, epigenomic, and transcriptomic data across brain and immune compartments highlights novel pathways linking autoimmunity with cognitive function and mental health. These insights offer a new conceptual framework for understanding comorbidity and suggest that targeting neuroimmune interfaces may benefit both metabolic and cognitive outcomes.

## Methods

### Study datasets and quality control

We used genome-wide association studies (GWAS) summary datasets and expression quantitative trait loci (eQTL) resources derived from individuals of predominantly European ancestry to minimize population heterogeneity and align with European linkage disequilibrium (LD) reference panels. The primary GWAS for T1D comprised 520,580 individuals (18,942 cases). Additional GWAS included cognitive traits (intelligence, educational attainment, executive function), psychiatric disorders (autism spectrum disorder, bipolar disorder, schizophrenia, obsessive-compulsive disorder, ADHD, major depression, PTSD, and neuroticism), neurological and neurodegenerative diseases (Alzheimer’s disease, Parkinson’s disease, amyotrophic lateral sclerosis, multiple sclerosis, myasthenia gravis, migraine, ischemic stroke), and sleep-related phenotypes (insomnia and sleep duration).

For regulatory analyses, we incorporated multiple eQTL datasets: bulk brain tissue from GTEx v8 (N=838), single-cell brain eQTLs spanning eight major cell types (N=192), single-cell blood eQTLs (N=982), and immune-cell eQTLs across 18 purified cell populations (N=200). These datasets enabled assessment of genetic regulation across both central nervous system and immune compartments.

Across all analyses, we applied stringent quality control procedures. SNPs were restricted to autosomal variants with minor allele frequency (MAF) >1% and imputation INFO >0.9. Variants with ambiguous strands were removed, and the extended major histocompatibility complex region (chr6:25–34 Mb) was excluded from analyses requiring LD independence due to complex long-range LD. Allele harmonization was performed across all datasets. LD estimation was based on the European reference panel from the 1000 Genomes Project Phase 3, ensuring ancestry-matched analyses.

This research complies with all the ethical regulations related to the secondary analysis of data collected by various cohorts, each of which obtained ethical approval and informed consent. All datasets used for analyses are summarized in **Supplementary Table 1**.

### Stratified LD Score Regression (S-LDSC)

We applied stratified LD score regression (S-LDSC) [32] to test whether T1D SNP-heritability was enriched in accessible chromatin regions of specific cortical cell types across human neurodevelopment. Single-nucleus ATAC-seq (snATAC-seq) peak annotations were obtained from cortex spanning prenatal, early postnatal (0–4 years), late postnatal (4–20 years), and adulthood (>20 years) [34]. Peaks were converted into binary annotation files for each cell type and stage. S-LDSC analyses were conducted using S-LDSC v1.0.1 with the baseline-LD model v2.2, applying European reference LD scores from 1000 Genomes Project Phase 3. Heritability enrichment was quantified as the proportion of heritability explained divided by the proportion of SNPs overlapping the annotation.

#### SCAVENGE analysis

SCAVENGE*—*Single Cell Analysis of Variant Enrichment through Network propagation of Genomic data—is a computational algorithm that uses network propagation to map causal variants to their relevant cellular context at single-cell resolution [36]. SCAVENGE mitigates sparsity in single-cell epigenomic data by leveraging network propagation: a small subset of “seed” cells enriched for trait-relevant variants are identified, and trait information is propagated across a nearest-neighbor cell graph to generate TRS for all cells. Following published procedures [36,37], we implemented SCAVENGE analysis to integrate fine-mapped genetic variants for T1D (n = 520,580 samples) [54] with single-nucleus ATAC-seq (snATAC-seq) profiles of human cortex across distinct developmental stages—prenatal, early postnatal (0–4 years), late postnatal (4–20 years), and adulthood (>20 years) [34]. This analysis yielded trait relevance scores (TRS) that quantify the contribution of each cell type and stage to T1D genetic predisposition.

To initiate propagation, fine-mapped T1D variants were intersected with accessible chromatin peaks to identify seed cells. Trait information from seed cells was propagated across a cell-to-cell similarity network, generating TRS for all nuclei. Cells with TRS > 2 were classified as enriched, consistent with the SCAVENGE framework [36]. To validate enrichment at the population level, TRS were aggregated across annotated cell types and developmental stages, and statistical significance was assessed using hypergeometric testing with false discovery rate (FDR) correction [37].

For benchmarking and cross-trait contextualization, the same workflow was applied using genome-wide significant loci from Alzheimer’s disease [55] and bipolar disorder [56] GWAS, enabling direct comparisons of developmental and cell-type-specific enrichment patterns across disorders. All SCAVENGE analyses were performed in R using the SCAVENGE software (v.1.0.2).

### Genome-wide genetic correlation analysis (cross-trait LDSC)

We applied cross-trait linkage disequilibrium score regression (LDSC) [57] to estimate genome-wide genetic correlations between T1D and each neurocognitive trait. Narrow-sense SNP-heritability and pairwise genetic correlations were computed from GWAS summary statistics for T1D and each neurocognitive phenotype. As previously described [57], LDSC estimates genetic correlation as the proportion of shared genetic variance between traits attributable to common SNP effects across the genome. Analyses were performed using LDSC v1.0.1 with pre-computed LD scores from European populations of the 1000 Genomes Project Phase 3. SNPs with minor allele frequency (MAF) > 0.05 were retained, while variants in the major histocompatibility complex (MHC, chr6:25–34 Mb) region were excluded due to extensive long-range LD.

To account for multiple testing across the 21 neurocognitive traits, we applied the Benjamini–Hochberg false discovery rate (FDR) procedure at q < 0.05. Both raw p-values and FDR-adjusted q-values are reported. Associations meeting q < 0.05 were considered statistically significant, whereas nominal associations (p < 0.05 but q ≥ 0.05) were reported as suggestive.

### Quantification of polygenic overlap by MiXeR analysis

We applied MiXeR [58], to quantify polygenic overlap between T1D and neurocognitive traits, providing a more nuanced measure of shared genetic architecture beyond global genetic correlation. MiXeR implements Gaussian causal mixture models to estimate both the number of causal variants unique to each trait and the number of variants with pleiotropic effects.

#### Univariate modeling

We first performed univariate MiXeR analysis (MiXeR v1.3) for T1D and each neurocognitive trait to estimate trait polygenicity (the number of SNPs accounting for 90% of SNP-heritability, *h*²_SNP_) and discoverability (the average effect size of trait-influencing variants). Variants with negligible effect sizes were excluded to ensure robust estimates.

#### Bivariate modeling

We then fit bivariate models to each T1D–neurocognitive trait pair to estimate the number of shared and unique causal variants. Model fit and convergence were evaluated using likelihood profiles and residual inspection. Dice coefficients were computed to summarize polygenic overlap on a 0–100% scale [59] and estimates of genome-wide genetic correlation were obtained using cross-trait LDSC for comparison.

#### Visualization

Results were summarized in Venn diagrams showing the number of unique and shared variants per trait pair. Conditional quantile–quantile (Q–Q) plots were generated to assess enrichment of SNP associations across traits [60]. Q–Q plots display the empirical cumulative distribution of P values in one phenotype, stratified by significance thresholds in the second phenotype (e.g., P ≤ 0.1, P ≤ 0.01, P ≤ 0.001), with leftward deflection from the null line indicating cross-trait enrichment.

#### Quality control

Analyses were restricted to autosomal SNPs with minor allele frequency (MAF) > 0.01 and imputation INFO score > 0.9, after removing strand-ambiguous and multi-allelic variants. The MHC region was excluded due to long-range linkage disequilibrium (LD). To ensure consistent LD structure across datasets, we used the European subset of the 1000 Genomes Project Phase 3 reference panel (n = 503) for LD estimation. All summary statistics were harmonized to the same genome build (GRCh37/hg19) with allele alignment performed against the reference panel.

The MiXeR analyses were conducted in R (v4.x) using the MiXeR v1.3 package with default settings unless otherwise specified.

### Shared locus and variant discovery using conjunctional FDR (conjFDR)

We applied conjunctional false discovery rate (conjFDR) analysis [38,39] to identify shared genetic loci influencing T1D and each neurocognitive phenotype. ConjFDR leverages cross-trait GWAS summary statistics to increase power for detecting pleiotropic variants while accounting for polygenic overlap.

As a first step, conditional quantile–quantile (Q–Q) plots were generated to evaluate enrichment of T1D-associated variants as a function of their significance in each neurocognitive phenotype (and vice versa). Evidence of leftward deflection in conditional Q– Q plots indicates polygenic overlap between the traits. Conditional FDR (condFDR) values were then computed for each SNP, representing the FDR of T1D associations given their significance in the neurocognitive phenotype (and vice versa). The conjFDR statistic was defined as the maximum of the two condFDR values per SNP, providing a conservative estimate of the probability that a variant is null for either or both traits. SNPs with conjFDR < 0.05 were considered jointly associated with T1D and the neurocognitive phenotype. Lead variants were annotated as the SNP with the lowest conjFDR value within each LD-independent locus, defined as r² < 0.1 within a ±250 kb window. LD was estimated using European reference data from the 1000 Genomes Project Phase 3. To minimize spurious associations, we used GWAS summary statistics pre-corrected for relevant covariates and population stratification. Analyses were restricted to autosomal SNPs, and only a single representative signal was retained in the extended major histocompatibility complex (MHC) region (chr6:26–34 Mb, hg19) due to complex LD.

All conjFDR analyses were conducted using the *pleioFDR* R package [60,61] following established workflows.

### Tissue enrichment analysis of shared loci between T1D and neurocognitive traits

We performed tissue enrichment analysis of loci jointly associated with T1D and neurocognitive traits as identified by conjFDR. SNPs with conjFDR < 0.05 were submitted to the Functional Mapping and Annotation (FUMA) platform (v1.5.2) [62] to define LD-independent loci. Independent significant SNPs were identified using the European 1000 Genomes Project Phase 3 reference panel [63], applying thresholds of r² < 0.6 for clumping and a maximum merging distance of ≤250 kb. Lead SNPs were defined as variants with r² < 0.1 within each locus, with the lead SNP taken as the variant with the lowest conjFDR value.

Novelty of lead SNPs was assessed by examining whether they achieved genome-wide significance (P < 5 × 10⁻⁸) in the original GWAS for T1D or the neurocognitive trait. Genes were assigned to loci by proximity, defined as the gene with the closest transcription start site to the lead SNP. Gene-level analyses were conducted using MAGMA within the FUMA framework, testing for tissue-specific expression and gene-set enrichment based on GTEx v8 transcriptomic data. Gene expression was quantified in transcripts per million (TPM). Multiple-testing correction was applied using FDR < 0.05. To further prioritize functional candidate genes, we integrated locus-to-gene assignments from Open Targets Genetics [64], focusing on lead SNPs outside of the extended MHC region due to its complex LD.

### Bidirectional Mendelian randomization analyses

We performed bidirectional two-sample Mendelian randomization (MR) to estimate causal relationships between T1D and neurocognitive phenotypes. MR uses genetic variants strongly associated with an exposure as instrumental variables (IVs) to test for potential causal effects of the exposure on the outcome.

#### Instrument selection and harmonization

SNPs associated with each exposure at genome-wide significance (P < 5 × 10⁻⁸; F statistic ≥ 10) were selected as IVs and pruned using LD clumping at r² < 0.001. Exposure and outcome summary statistics were harmonized by aligning alleles to the forward strand; ambiguous SNPs with non-inferable strands were excluded.

#### Primary MR analyses

SNP-specific causal effects were estimated using the Wald ratio and meta-analyzed across variants with the inverse-variance weighted (IVW) method under a multiplicative random-effects model [65]. IVW was used as the primary MR test, under the assumption that all IVs are valid and horizontal pleiotropy is absent.

#### Sensitivity analyses

To evaluate robustness to horizontal pleiotropy, we performed MR using the weighted median (WM) and MR-Egger regression methods [65–68]. The weighted median estimator provides consistent causal effect estimates if at least 50% of the weight comes from valid IVs, even if the remaining IVs are invalid. MR-Egger allows for directional pleiotropy by including an intercept term in the regression of outcome coefficients on exposure coefficients; the intercept test was used to evaluate the presence of unbalanced pleiotropy [67–69]. Under the null of no pleiotropy, the MR-Egger intercept approaches zero, and MR-Egger converges to IVW [65,70].

#### Multiple testing correction

To control for the number of MR tests conducted, we applied the Benjamini–Hochberg FDR procedure across all 42 primary IVW exposure–outcome pairs (21 neurocognitive traits tested bidirectionally). Associations meeting q < 0.05 were considered statistically significant, whereas those with p < 0.05 but q ≥ 0.05 were reported as nominal. Weighted median and MR-Egger estimates were treated as sensitivity analyses and were not subjected to further multiplicity correction.

#### Reporting

Causal effect estimates are reported as odds ratios (OR) with 95% confidence intervals (CI), raw p-values, and FDR-adjusted q-values for the outcome per one standard deviation (SD) increase in the exposure.

All analyses were conducted in R (v4.3.2; R Foundation for Statistical Computing, Vienna, Austria) using the *TwoSampleMR* package. Methods and reporting adhered to the STROBE-MR best-practice guidelines [71].

### Summary-based Mendelian randomization and heterogeneity in dependent instruments (SMR/HEIDI) analysis

SMR extends the concept of MR, allowing testing of the hypothesis that genetically determined expression levels of a gene are associated with a phenotype. A key assumption of this approach is that the same underlying causal variant determines both gene expression and the disease/trait. SMR cannot distinguish between vertical pleiotropy—the situation in which variant influences phenotype *via* gene expression, and horizontal pleiotropy, the situation in which variant influences phenotype and gene expression but influences the phenotype at least partly independently of gene expression. To distinguish pleiotropy from linkage, the heterogeneity in dependent instruments (HEIDI) test was developed [53]. which exploits the observation that if gene expression and disease/trait are in vertical pleiotropy with the same causal variant, the causal effect is identical for any variant in linkage disequilibrium (LD) with the causal variant. Thus, greater heterogeneity among effect size statistics calculated for all significant *cis*-eQTLs implies a greater likelihood that linkage, rather than causality/vertical pleiotropy, explains the observed causal association. The heterogeneity statistic, the ‘HEIDI’ statistic, tests the hypothesis HEIDI = 0. This provides a formal test of heterogeneity, with *P*-values < 0.05 suggestive of linkage, rather than pleiotropy, as the underlying biological model [53]. Thus, the SMR & HEIDI approach is widely used to test if a transcript and phenotype are associated because of a shared causal variant (i.e., pleiotropy). We used SMR/HEIDI to integrate GWAS association signals for T1D and each neurocognitive trait with eQTL derived from bulk brain regions [72], and single-nucleus brain cells [73] and single-cell blood [74] and immune cells [75] to identify genes whose expression levels are jointly associated with T1D and neurocognitive traits. We performed SMR using the SMR software tool (SMR v1.0.2) in the command line using default options [53], i.e., *cis*-eQTLs selected based on minimum *P* = 5 × 10^−8^, eQTLs included for the HEIDI test based on minimum *P* = 1.57 × 10^−3^, eQTLs included for the HEIDI test if *R*^2^ with the top *cis*-eQTL was between 0.05 and 0.9, minimum number of SNPs included in the HEIDI test = 3, maximum number of SNPs included in the HEIDI test = 20 and physical window around probe within which the top *cis*-eQTL was selected = 2 MB. *P*-values were adjusted in R (v3.6.1) to control the false discovery rate at *ɑ* = 0.05 using the Benjamini–Hochberg procedure. Associations with *P*_HEIDI_<0.05—cutoff [76]—were considered likely due to linkage. Probes were excluded if any of the transcript or the top eQTL resided within the super-extended major histocompatibility complex (hg19 6:25 000 000–35 000 000) given the complex linkage disequilibrium structures within this region. Linkage disequilibrium estimation was performed using reference genomes obtained from the 1000 genomes samples of European ancestry [77]).

### Transcriptomic and proteomic analysis for differential expression of SMR/HEIDI-identified genes in Parkinson’s disease-affected and control brain tissue

To investigate whether genetically predicted expression of genes in the brain associated with T1D manifests in human disease contexts, we analyzed our previously published single-nucleus transcriptomic and proteomic data from brain tissue of individuals with Parkinson’s disease (PD; *n* = 6) and matched control individuals (*n* = 6) [47]. This focus was motivated by suggestive Mendelian randomization (MR) associations between T1D and PD— particularly the nominal evidence for increased T1D risk with PD liability—together with conjFDR and SMR/HEIDI analyses identifying pleiotropic expression quantitative trait loci (eQTLs) shared between the two traits. We interrogated our previously published single-nucleus RNA-sequencing (snRNA-seq) and label-free quantitative proteomics datasets generated from the prefrontal cortex of PD patients and control subjects (PMID: 39475571). Genes were selected for transcriptomic and proteomic interrogation if their genetically predicted expression in bulk brain tissue (cortex, frontal cortex BA9, substantia nigra) or microglia was significantly associated with T1D risk based on SMR/HEIDI analysis.

Transcript-level differential expression analyses were conducted using the DESeq2 framework, incorporating covariate adjustment for age, sex, and postmortem interval, and following the analytical pipeline detailed in our previous publication. Genes with an adjusted P < 0.05 were considered differentially expressed.

In parallel, we assessed differential protein abundance of SMR-identified genes using label-free mass spectrometry-based proteomics data from the same cohort. Protein-level comparisons were performed using moderated t-tests with Benjamini–Hochberg false discovery rate correction. Given the modest sample size and resulting limited statistical power, we adopted a relaxed significance threshold of P < 0.1 to capture a broader set of biologically relevant signals.

## Data availability

Genome-wide association study (GWAS) summary statistics used in this study are publicly available from the original consortia and repositories listed in Supplementary Table 1, with accession links and/or PMIDs provided. Single-cell ATAC-seq and RNA-seq data are available from the referenced studies as indicated in the Methods. Processed data generated in this work, including results from LDSC, MiXeR, conjFDR, SMR/HEIDI, and Mendelian randomization analyses, are provided in the supplementary tables.

## Code availability

All analysis scripts and pipelines used in this study are openly available at: https://github.com/AlagsLabTeam/T1D-NEURO

## Acknowledgements

This study was supported by Breakthrough T1D grants to DAA (Grant Keys: 5-CDA-2025-1682-S-B and SRA-2024-1472-S-B).

## Author contributions

PS and ZAS contributed to the conception and initiation of the project. PS, ZAS, ZX, YD, and BZ performed data analyses. DAA conceived, designed, and supervised the study and drafted the manuscript. All authors (PS, ZAS, ZX, YD, AJ, MS, SR, BZ, LZ, ATD, SA, and DAA) contributed to data interpretation, manuscript review, and approval of the final draft.

## Competing interests

All authors declare no competing interests.

## Notes

### Competing Interest Statement

The authors have declared no competing interest.

### Author Declarations

This study analyzed only publicly available, de-identified GWAS and eQTL summary statistics. No individual-level human data were accessed, and therefore ethical approval was not required.

### Summary of Updates

This version corrects a duplicated figure legend. No results, analyses, or conclusions have been changed.

