## Supplementary Table 1 for "Shared genetic and neuroimmune architecture links type 1 diabetes with neurocognitive traits"

**Table 1: GWAS and eQTL summary datasets used for analyses.**

| No. | Trait / Dataset | Population | Sample size | PMID | Accession No/Reference |
| --- | --- | --- | --- | --- | --- |
| 1 | T1D | European | 520 580 (18 942 cases) | 34012112 | GCST90014023 |
| 2 | Bipolar disorder | European | 413 466 (41 917 cases) | 34002096 |  |
| 3 | ASD | European | 46 350 (18 381 cases) | 30804558 |  |
| 4 | Alzheimer’s disease | European | 487 511 (85 934 cases) | 35379992 | GCST90027158 |
| 5 | Parkinson’s disease | European | 1 019 060 (37 688 cases) | 31701892 | GCST009324 |
| 6 | Multiple sclerosis | European | 38 589 (14 498 cases) | 24076602 | GCST005531 |
| 7 | Myasthenia gravis | European | 38 243 (1 873 cases) | 35074870 | GCST90093061 |
| 8 | Major depression | European | 92 957 (29 475 cases) | 31969693 | GCST009979 |
| 9 | Intelligence | European | 269 867 | 29942086 | GCST006250 |
| 10 | Education | European | 405 072 | 27225129 | GCST003676 |
| 11 | Ischemic Stroke | European | 1, 296, 908 (62, 100 cases) | 36180795 | GCST90104540 |
| 12 | OCD | European | 9 725 (2 688 cases) | 28761083 |  |
| 13 | Migraine | European | 513 266 (26 052 cases) | 37415806 | GCST90271641 |
| 14 | Schizophrenia | European | 130 644 (53 386 cases) | 35396580 |  |
| 15 | Insomnia | European | 2 365 010 (593 724 cases) | 35835914 |  |
| 16 | Sleep duration | European | 445 966 | 37770476 |  |
| 17 | ADHD | European | 225 534 (38 691 cases) | 36702997 |  |
| 18 | Neuroticism | European | 393 411 | 29892013 | GCST90029028 |
| 19 | PTSD | European | 1 222 882 (137 136 cases) | **38637617** |  |
| 20 | ALS | European | 138 086 (27 205 cases) | 34873335 | GCST90027164 |
| 21 | Executive function | European | 427 037 | 36150907 | GCST90162547 |
| 22 | SC-blood eQTL | European | 982 | 35389779 | [1] |
| 23 | Regional brain eQTL | European | 838 (GTEx v8) | **29022597** | [2] |
| 24 | SC-brain eQTL | European | 192 | 35915177 | [3] |
| 25 | 18 immune cell types | European | 200 | **27863251** |  |

**Supplementary Table 1 | GWAS and eQTL datasets used in this study.**
Summary genome-wide association studies (GWAS) and expression quantitative trait loci (eQTL) resources included in the analyses. All datasets were derived from individuals of predominantly European ancestry. GWAS covered T1D, cognitive and educational traits, psychiatric disorders, neurological diseases, and sleep phenotypes, with sample sizes ranging from tens of thousands to over two million participants. eQTL resources included bulk brain regions (GTEx v8), single-cell brain and blood, and purified immune cell types. For all datasets, we applied consistent quality control filters (autosomal SNPs, MAF >1%, INFO >0.9, allele harmonization, exclusion of ambiguous strands, and removal of the extended MHC region). Linkage disequilibrium was estimated using European reference panels from the 1000 Genomes Project Phase 3. T1D=type 1 diabetes; EXF= executive functioning; Sc= single cell; eQTL= expression quantitative trait loci; CHIP= clonal hematopoiesis of indeterminate potential; ASD= autism spectrum disorders; AD= Alzheimer’s disease; PD= Parkinson’s disease; MS= multiple sclerosis; MG= myasthenia gravis; OCD= obsessive-compulsive disorder; ASP= antisocial personality disorder; ADHD= attention deficit hyperactive disorder; PTSD= post-traumatic stress disorder; ALS= amyotrophic lateral sclerosis; EDA= educational attainment; SC-blood eQTL= single-cell blood eQTL; SC-brain eQTL= single-cell brain eQTL.
