## Supplementary Table 5 for "Shared genetic and neuroimmune architecture links type 1 diabetes with neurocognitive traits"

| eQTL | Chr | Gene | Tissue/Cell | β_eQTL_ | *P*_eQTL_ | β_T1D_ | *P*T1D | Neuro | βNeuro | *P*Neuro |
| --- | --- | --- | --- | --- | --- | --- | --- | --- | --- | --- |
| rs62061822 | 17 | CRHR1-IT1 | Substantia nigra | 1.0734 | 8.69E-27 | -0.086621 | 1.63E-06 | ASD | 0.0717345 | 3.62E-05 |
| rs58879558 | 17 | LRRC37A4P | Substantia nigra | -1.331 | 4.40E-29 | 0.0701564 | 1.62E-06 | ASD | -0.0581891 | 2.42E-05 |
| rs2838 | 17 | CRHR1-IT1 | Spinal cord cervical C1 | 1.27962 | 1.82E-35 | -0.07385 | 9.97E-07 | ASD | 0.0597858 | 2.82E-05 |
| rs62054822 | 17 | LRRC37A4P | Spinal cord cervical C1 | -1.2359 | 9.82E-33 | 0.0744344 | 1.16E-06 | ASD | -0.0625464 | 2.37E-05 |
| rs8073146 | 17 | LRRC37A2 | Hypothalamus | 1.30903 | 1.56E-43 | -0.070381 | 6.77E-06 | ASD | 0.0601185 | 1.40E-05 |
| rs62057151 | 17 | LRRC37A4P | Frontal cortex BA9 | -1.194 | 1.55E-47 | 0.0741837 | 1.13E-06 | ASD | -0.065157 | 1.42E-05 |
| rs2532343 | 17 | RP11-259G18.1 | Cerebellum | 1.23481 | 8.89E-51 | -0.075679 | 4.51E-07 | ASD | 0.0647073 | 9.11E-06 |
| rs1131017 | 12 | RPS26 | Substantia nigra | -1.0664 | 8.19E-36 | 0.214647 | 5.10E-23 | EDU | 0.013128 | 1.23E-05 |
| rs111511018 | 17 | RP11-259G18.3 | Putamen basal ganglia | 1.25967 | 5.53E-38 | -0.075028 | 8.66E-07 | EDU | -0.0119079 | 3.14E-06 |
| rs10876864 | 12 | SUOX | Nuc accum basal ganglia | 0.377372 | 2.47E-11 | -0.601126 | 7.07E-10 | EDU | -0.0397486 | 6.12E-07 |
| rs240704 | 16 | NPIPB7 | Nuc accum basal ganglia | -0.54205 | 1.88E-12 | 0.205599 | 1.79E-07 | EDU | 0.0239828 | 1.78E-06 |
| rs1131017 | 12 | RPS26 | hypothalamus | -0.99493 | 4.36E-54 | 0.23007 | 5.25E-29 | EDU | 0.0140713 | 7.89E-06 |
| rs8073146 | 17 | LRRC37A2 | hypothalamus | 1.30903 | 1.56E-43 | -0.070381 | 6.77E-07 | EDU | -0.0129867 | 1.57E-07 |
| rs112560196 | 17 | CRHR1-IT1 | Anterior cingulate cortex | 0.768988 | 7.69E-30 | -0.076677 | 3.55E-06 | EDU | -0.020806 | 1.39E-06 |
| rs8073146 | 17 | LRRC37A2 | Caudate Basal ganglia | 1.22712 | 4.29E-42 | -0.075079 | 7.18E-07 | EDU | -0.0138536 | 1.69E-07 |
| rs2942166 | 17 | LRRC37A2 | Amygdala | 1.09695 | 2.66E-28 | -0.086798 | 9.59E-07 | EXF | -0.0071561 | 2.64E-07 |
| rs55974014 | 17 | DND1P1 | Amygdala | 1.04281 | 1.35E-19 | -0.086937 | 7.94E-06 | EXF | -0.007885 | 5.42E-07 |
| rs55974014 | 17 | RP11-259G18.3 | Amygdala | 1.18792 | 4.05E-27 | -0.076317 | 3.55E-06 | EXF | -0.0069221 | 1.49E-07 |
| rs79172804 | 17 | PLEKHM1 | Caudate Basal ganglia | 0.399783 | 2.31E-11 | -0.268566 | 1.36E-05 | EXF | -0.0231669 | 4.02E-06 |
| rs62056905 | 17 | CRHR1-IT1 | Caudate Basal ganglia | 0.886044 | 1.06E-38 | -0.104362 | 7.81E-07 | EXF | -0.008799 | 1.32E-07 |
| rs56340658 | 17 | PLEKHM1 | Cerebellar hemisphere | -0.84891 | 1.18E-29 | 0.116634 | 3.55E-07 | EXF | 0.00917653 | 2.75E-07 |
| rs55974014 | 17 | KANSL1-AS1 | Cerebellar hemisphere | 1.17298 | 8.29E-37 | -0.077289 | 1.94E-06 | EXF | -0.0070103 | 5.50E-08 |
| rs56046792 | 17 | ARL17A | Cerebellar hemisphere | 1.1204 | 6.49E-34 | -0.083534 | 7.98E-07 | EXF | -0.0069673 | 1.77E-07 |
| rs55974014 | 17 | FAM215B | Cerebellum | 0.873863 | 6.29E-24 | -0.103745 | 4.73E-06 | EXF | -0.0094098 | 2.38E-07 |
| rs55974014 | 17 | ARL17A | Cerebellum | 1.13378 | 4.39E-38 | -0.079961 | 1.83E-06 | EXF | -0.0072527 | 4.98E-08 |
| rs55974014 | 17 | KANSL1-AS1 | Cerebellum | 1.1736 | 2.14E-44 | -0.077248 | 1.44E-06 | EXF | -0.0070066 | 3.29E-08 |
| rs17573607 | 17 | SPPL2C | Cerebellum | 0.758482 | 6.65E-25 | -0.124658 | 1.45E-06 | EXF | -0.010388 | 3.78E-07 |
| rs62055937 | 17 | MAPT-AS1 | Nuc accum basal ganglia | -0.72614 | 1.07E-21 | 0.127806 | 3.32E-06 | EXF | 0.0106645 | 9.03E-07 |
| rs2942166 | 17 | LRRC37A2 | Amygdala | 1.18792 | 4.05E-27 | -0.086798 | 9.59E-07 | INT | -0.0151999 | 7.01E-06 |
| rs55974014 | 17 | DND1P1 | Amygdala | 1.04281 | 1.35E-19 | -0.086937 | 7.94E-06 | INT | -0.0159296 | 2.55E-05 |
| rs62034350 | 16 | SULT1A1 | Caudate basal ganglia | -0.52776 | 2.68E-09 | 0.186782 | 8.10E-06 | INT | 0.0485439 | 7.24E-07 |
| rs56046792 | 17 | ARL17A | Cerebellar hemisphere | 1.1204 | 6.49E-34 | -0.083534 | 7.98E-07 | INT | -0.0150707 | 4.57E-06 |
| rs62055937 | 17 | MAPT-AS1 | Nuc accum basal ganglia | -0.72614 | 1.07E-21 | 0.127806 | 3.32E-06 | INT | 0.0231597 | 1.21E-05 |
| rs58879558 | 17 | LRRC37A4P | Substantia nigra | -1.33181 | 4.40E-29 | 0.0701564 | 1.62E-06 | INT | 0.0111979 | 3.92E-05 |
| rs2942166 | 17 | LRRC37A2 | Amygdala | 1.09695 | 2.66E-28 | -0.086798 | 9.59E-07 | PD | -0.198004 | 1.27E-08 |
| rs55974014 | 17 | DND1P1 | Amygdala | 1.04281 | 1.35E-19 | -0.086937 | 7.94E-06 | PD | -0.206653 | 8.29E-08 |
| rs393152 | 17 | ARL17A | Anterior cingulate cortex | 0.88759 | 1.91E-14 | -0.104202 | 1.20E-05 | PD | -0.23085 | 8.22E-07 |
| rs62056905 | 17 | CRHR1-IT1 | Caudate basal ganglia | 0.886044 | 1.06E-38 | -0.104362 | 7.81E-07 | PD | -0.236783 | 4.37E-09 |
| rs55974014 | 17 | KANSL1-AS1 | Cerebellar hemisphere | 1.17298 | 8.29E-37 | -0.077289 | 1.94E-06 | PD | -0.18372 | 3.88E-09 |
| rs10445367 | 17 | PLEKHM1 | Cortex | 0.491111 | 2.89E-16 | -0.213929 | 5.24E-06 | PD | -0.424344 | 1.94E-06 |
| rs373695612 | 17 | KANSL1-AS1 | Naïve B cells | 1.03814 | 1.47E-14 | -0.086841 | 3.27E-05 | EXF | -0.0077236 | 4.00E-06 |
| rs62062288 | 17 | LRRC37A2 | Naïve B cells | 1.09489 | 1.17E-12 | -0.084248 | 2.63E-05 | EXF | -0.0073083 | 6.66E-06 |
| rs56040418 | 17 | LRRC37A2 | Naïve CD4+ T cells | 1.03094 | 4.71E-11 | -0.090461 | 3.03E-05 | EXF | -0.0077397 | 1.08E-05 |
| rs1560312 | 17 | CRHR1-IT1 | Naïve CD4+ T cells | 1.05263 | 3.95E-12 | 0.0179344 | 2.52E-05 | EXF | 0.0014544 | 1.33E-05 |
| rs373695612 | 17 | KANSL1-AS1 | Naïve CD4+ T cells | 1.17554 | 2.44E-15 | -0.07669 | 2.81E-05 | EXF | -0.0068209 | 3.18E-06 |
| rs2732712 | 17 | LRRC37A4P | Naïve CD4+ T cells | -1.23278 | 2.00E-18 | 0.074226 | 7.31E-06 | EXF | 0.00665551 | 7.98E-07 |
| rs2532387 | 17 | DND1P1 | Stimulated CD4+ T | 1.19687 | 7.94E-15 | -0.0784 | 9.57E-06 | EXF | -0.0066045 | 3.73E-06 |
| rs8076131 | 17 | GSDMB | Stimulated CD4+ T | 0.967513 | 4.46E-11 | -0.077761 | 3.98E-05 | MG | -0.12 | 2.41E-05 |
| rs2532387 | 17 | KANSL1-AS1 | Naïve CD8+ T cells | 1.15471 | 1.74E-15 | -0.081262 | 8.15E-06 | EXF | -0.0068456 | 3.09E-06 |
| rs56040418 | 17 | LRRC37A2 | Naïve CD8+ T cells | 1.12002 | 7.87E-14 | -0.083267 | 1.22E-05 | EXF | -0.0071241 | 3.49E-06 |
| rs9388490 | 6 | CENPW | Stimulated CD8+ T | -0.86736 | 2.60E-10 | -0.120165 | 1.50E-06 | EDU | -0.0126821 | 3.34E-05 |
| rs9388490 | 6 | CENPW | Stimulated CD8+ T | -0.86736 | 2.60E-10 | -0.120165 | 1.50E-06 | INT | -0.0166224 | 5.26E-05 |
| rs753235 | 17 | CRHR1-IT1 | Monocytes | 1.22447 | 2.15E-19 | -0.075802 | 3.92E-06 | EXF | -0.00655 | 6.90E-07 |
| rs373695612 | 17 | KANSL1-AS1 | Monocytes | 1.22085 | 2.07E-16 | -0.073844 | 2.32E-05 | EXF | -0.0065677 | 2.38E-06 |
| rs11102702 | 1 | AP4B1 | QT monocytes | -0.71549 | 5.53E-11 | -0.494407 | 4.63E-10 | MG | -0.275191 | 1.09E-06 |
| rs112010353 | 17 | KANSL1-AS1 | QT Neutrophils | 1.29705 | 1.31E-50 | -0.074398 | 2.54E-07 | EXF | -0.0059313 | 1.08E-07 |
| rs113208333 | 16 | SULT1A1 | QT CD4+ T cells | -0.59297 | 3.28E-11 | 0.178308 | 2.02E-06 | INT | 0.0423928 | 1.13E-07 |
| rs71624119 | 5 | ANKRD55 | QT CD4+ T cells | -1.06081 | 3.27E-22 | 0.0601332 | 1.10E-05 | MS | 0.110171 | 3.98E-07 |
| rs11171739 | 12 | ERBB3 | TH1 cells | 0.983791 | 2.11E-10 | -0.225021 | 3.86E-09 | INT | -0.014699 | 5.27E-05 |
| rs705705 | 12 | SUOX | Memory Treg | -1.23473 | 8.76E-15 | -0.174038 | 7.69E-06 | EDU | -0.0145781 | 2.08E-06 |
| rs705705 | 12 | SUOX | Memory Treg | -1.23473 | 8.76E-15 | -0.174038 | 7.69E-06 | INT | -0.0121049 | 2.06E-05 |
| rs2640564 | 12 | SUOX | Naïve Treg | -1.21829 | 8.34E-14 | -0.19956 | 9.83E-12 | EDU | -0.0147749 | 2.92E-06 |
| rs2640564 | 12 | SUOX | Naïve Treg | -1.21829 | 8.34E-14 | -0.19956 | 9.83E-12 | INT |  |  |
| rs62062288 | 17 | RP11-156P1.2 | Astrocytes | 0.855133 | 1.79E-20 | -0.10787 | 5.54E-06 | ASD | 0.09402 | 4.17E-05 |
| rs70602 | 17 | KANSL1 | Endothelial cells | 0.948218 | 1.13E-15 | -0.078786 | 2.45E-04 | ASD | 0.0786709 | 1.68E-04 |
| rs70602 | 17 | KANSL1 | Endothelial cells | 0.948218 | 1.13E-15 | -0.078786 | 2.45E-04 | EXF | -0.0078464 | 6.93E-06 |
| rs70602 | 17 | KANSL1 | Endothelial cells | 0.948218 | 1.13E-15 | -0.078786 | 2.45E-04 | INT | -0.0159903 | 1.17E-04 |
| rs2668622 | 17 | KANSL1 | Excitatory neurons | 1.17235 | 1.12E-24 | -0.07819 | 4.90E-06 | ASD | 0.0660179 | 4.12E-05 |
| rs2668622 | 17 | KANSL1 | Excitatory neurons | 1.17235 | 1.12E-24 | -0.07819 | 4.90E-06 | EXF | -0.0068699 | 3.83E-07 |
| rs2668622 | 17 | KANSL1 | Excitatory neurons | 1.17235 | 1.12E-24 | -0.07819 | 4.90E-06 | PD | -0.180748 | 4.02E-08 |
| rs7210219 | 17 | RP11-105N13.4 | Excitatory neurons | 0.572624 | 5.25E-22 | -0.166841 | 2.12E-06 | EXF | -0.0162445 | 8.86E-06 |
| rs2668622 | 17 | ARL17B | Inhibitory neurons | 1.3714 | 5.63E-28 | -0.066841 | 3.71E-06 | ASD | 0.0564359 | 3.44E-05 |
| rs1991556 | 17 | LRRC37A2 | Inhibitory neurons | 1.00987 | 3.72E-19 | -0.089155 | 7.37E-06 | ASD | 0.0739691 | 9.35E-05 |
| rs2668622 | 17 | ARL17B | Inhibitory neurons | 1.3714 | 5.63E-28 | -0.066841 | 3.71E-06 | EXF | -0.0058728 | 2.51E-07 |
| rs1991556 | 17 | LRRC37A2 | Inhibitory neurons | 1.00987 | 3.72E-19 | -0.089155 | 7.37E-06 | INT | -0.0157692 | 4.33E-05 |
| rs2668622 | 17 | ARL17B | Inhibitory neurons | 1.3714 | 5.63E-28 | -0.066841 | 3.71E-06 | INT | -0.0121555 | 1.14E-05 |
| rs1991556 | 17 | LRRC37A2 | Inhibitory neurons | 1.00987 | 3.72E-19 | -0.089155 | 7.37E-06 | PD | -0.190916 | 9.51E-08 |
| rs2668622 | 17 | ARL17B | Inhibitory neurons | 1.3714 | 5.63E-28 | -0.066841 | 3.71E-06 | PD | -0.154514 | 2.25E-08 |
| rs2668622 | 17 | ARL17B | Microglia | 1.35237 | 3.70E-30 | -0.067782 | 3.18E-06 | ASD | 0.05723 | 3.11E-05 |
| rs2668622 | 17 | ARL17B | Microglia | 1.35237 | 3.70E-30 | -0.067782 | 3.18E-06 | EXF | -0.0059554 | 1.98E-07 |
| rs2668622 | 17 | ARL17B | Microglia | 1.35237 | 3.70E-30 | -0.067782 | 3.18E-06 | INT | -0.0123265 | 1.01E-05 |
| rs2668622 | 17 | ARL17B | Microglia | 1.35237 | 3.70E-30 | -0.067782 | 3.18E-06 | PD | -0.156688 | 1.62E-08 |
| rs55915917 | 17 | ARL17B | Perivascular vessel | 1.47437 | 4.23E-30 | -0.063171 | 1.85E-06 | ASD | 0.0556847 | 1.74E-05 |
| rs55915917 | 17 | ARL17B | Perivascular vessel | 1.47437 | 4.23E-30 | -0.063171 | 1.85E-06 | EDU | -0.0115303 | 3.89E-07 |
| rs62062288 | 17 | KANSL1 | Pericytes | 1.16619 | 9.33E-17 | -0.079097 | 1.01E-05 | ASD | 0.0689421 | 6.24E-05 |
| rs62062288 | 17 | KANSL1 | Pericytes | 1.16619 | 9.33E-17 | -0.079097 | 1.01E-05 | Migraine | 0.027093 | 6.18E-04 |
| rs2532233 | 17 | KANSL1-AS1 | CD4 T cells | 0.7478 | 2.65E-22 | -0.125015 | 2.58E-06 | ASD | 0.106046 | 3.11E-05 |
| rs2532233 | 17 | KANSL1-AS1 | CD4 T cells | 0.7478 | 2.65E-22 | -0.125015 | 2.58E-06 | EDU | -0.0187216 | 2.60E-05 |
| rs72793812 | 16 | TFUM | CD4 T cells | 0.6072 | 5.72E-13 | -0.128496 | 1.61E-05 | EDU | -0.0230567 | 8.96E-05 |
| rs62037364 | 16 | TFUM | CD8 T cells | 0.4613 | 7.00E-10 | -0.162359 | 7.43E-05 | EDU | -0.030349 | 1.98E-04 |
| rs7297175 | 12 | RPS26 | DCs | -0.9141 | 2.77E-39 | 0.243222 | 7.78E-24 | EDU | 0.0153156 | 1.10E-05 |
| rs7297175 | 12 | RPS26 | DCs | -0.9141 | 2.77E-39 | 0.243222 | 7.78E-24 | EXF | 0.00474771 | 4.75E-04 |
| rs7297175 | 12 | RPS26 | DCs | -0.9141 | 2.77E-39 | 0.243222 | 7.78E-24 | INT | 0.0157311 | 1.58E-06 |
| rs62037364 | 16 | TFUM | DCs | 0.3889 | 2.66E-08 | -0.192584 | 1.52E-04 | INT | -0.0705806 | 1.40E-06 |
| rs7297175 | 12 | RPS26 | DCs | -0.9141 | 2.77E-39 | 0.243222 | 7.78E-24 | MS | -0.0574962 | 2.22E-03 |

Nuc. accum. Basal ganglia= Nucleus accumbens basal ganglia
